# Pre- and post-operative psychological interventions to prevent pain and fatigue after breast cancer surgery (PREVENT): a randomized controlled trial

**DOI:** 10.1101/2022.05.05.22274733

**Authors:** Silje E. Reme, Alice Munk, Marianne Therese Smogeli Holter, Ragnhild S. Falk, Henrik. B. Jacobsen

## Abstract

**Background:** Breast cancer is the most common cancer type among women worldwide with over a million new cases each year. More than 40% of these women will struggle with chronic pain and fatigue after surgery, regardless of surgical procedure. These consequences are detrimental and result in distress and disability, including work disability. Few attempts have been made to prevent chronic pain and fatigue after surgery by applying a psychological approach, despite psychological risk factors being crucial in the development of both chronic pain and fatigue. In this study, we aim to develop and test an easily implementable strategy of preventing chronic pain and fatigue after breast cancer surgery. The intervention strategy involves a pre-operative hypnosis session and a web-based post-operative Acceptance and Commitment Therapy (ACT). The hypnosis has previously been found effective in alleviating acute post-operative pain and fatigue in breast cancer patients, while ACT is well suited to cancer populations as it offers a model of healthy adaptation to difficult circumstances. Together they form an intervention strategy with both a preventive and a rehabilitative focus.

**Methods/design:** This randomized controlled trial aims to estimate the effects of the pre- and post-operative interventions compared to attentional control and treatment as usual (TAU) and will also include a qualitative process evaluation. Participants will be randomized to receive either a pre-operative brief hypnosis session and a post-operative web-based psychological intervention (iACT) or a pre-operative one-session mindfulness through an audio file and post-operative TAU. Self-reported questionnaire data and biomarker data will be assessed pre-surgery, post-surgery and 3 and 12 months after surgery. In addition, we will assess registry data on sick leave and prescriptions until 2-year follow-up. In the qualitative process evaluation, data will be collected from participants from both study arms (through interviews and a diary) and two different analyses performed (socio-narrative and Grounded Theory) with the objective to describe the development of chronic post-surgical pain and fatigue and the potential influence of the interventions on these processes. The study is set-up to demonstrate a minimum difference in pain of 1 point on NRS (0-10) and 3 points on FACIT-F (0-52) between the groups at 3-months follow-up by including 200 breast cancer patients in total.

**Discussion:** This trial will be the first study to estimate the effect of a combined pre-operative hypnosis with a post-operative iACT to prevent pain and fatigue after breast cancer surgery. The results from our study might i) help the large group of women affected by chronic pain and fatigue after breast cancer surgery, ii) shed light on the mechanisms involved in chronic pain and fatigue development, and iii) serve as a model for other surgical procedures.

**Trial registration:** Clinicaltrials.gov, registration number NCT04518085. Registered on January 29^th^, 2020.

## Background

### Late effects from breast cancer

Breast cancer is the most prevalent cancer type among women, with >1 million new cases worldwide every year (1). Survival rates are increasing (2), but for many patients survivorship is also characterized by disabling psychological and physical late effects (3–5).

Chronic post-surgical pain (CPSP) and chronic fatigue are most prevalent. CPSP has prevalence rates ranging from 25-60% depending on definition, measurement and treatment (6). In a recent Norwegian study, more than 40% of women treated for breast cancer continued to have pain 2-6 years after surgery (7), while over a third of the women in a large Danish study reported chronic pain 5-7 years after surgery (8).

### Chronic fatigue after surgery

Chronic fatigue is the most common symptom associated with cancer and its treatments (9, 10), and is usually operationalized as fatigue lasting 6 months or longer (11). Despite a high prevalence of chronic fatigue, studies on the occurrence of and predictors of fatigue after surgery are scarce. One of the few studies investigating chronic fatigue after breast cancer surgery reported high levels of fatigue in the first two months after surgery, followed by mild-to-moderate levels of fatigue that persisted 12 months after surgery (12). In another study of breast cancer survivors, a third of the women struggled with chronic fatigue up to 10 years into survivorship (13).

### Chronic pain after surgery

Surgery as a major risk factor for chronic pain was first identified in 1998 (14), but has since received increasing attention and priority (15). Chronic post-surgical pain has been defined as pain that persists at least 3 months after surgery, localized to the surgical site or referred area, and not explained by other causes (16, 17). The pathophysiology of CPSP is still unclear. Presumably, the nature of most CPSP is neuropathic since surgery (e.g., mastectomy) may involve major nerve damage, and is associated with the highest incidence of such pain (16, 18). However, many patients with CPSP do not show any signs of neuropathic pain, or any sensory changes (3). It is thus assumed that the distinct pathophysiology reflects both peripheral and central sensitization as well as humoral factors contributing to pain (19), and CPSP is now grounded in the bio-psycho-social model, where psychological and social factors contribute to biological factors in the development of chronic pain (20, 21). Interestingly, there are several shared risk factors between pain and fatigue after surgery, including depression and outcome expectancies (22), implying that the very same processes might influence both post-surgical pain and fatigue.

### Risk factors

Documented risk factors for CPSP can be divided into more or less modifiable factors. Female gender, younger age (18) and preoperative pain conditions (16, 18, 21, 23) are the most commonly reported unmodifiable risk factors, along with pre-operative use of opioids (21) which is also on the less modifiable side. However, a preventive intervention could target psychological factors, which are both modifiable, robust, and potent predictors of CPSP and fatigue (20, 24). The most frequent tap into anxiety, such as hypervigilance and pain catastrophizing (24–28). Other significant predictors involve depression, stress, and optimism (12, 20, 27).

### Theoretical model

Our research group recently developed a theoretical model for CPSP following breast cancer surgery (the surgery outcome expectancy model of CPSP, “SURGE”) (19). The key principles in the SURGE model builds on the cognitive activation theory of stress (29), predictive coding accounts (30), and well-established psychoneuroimmunological processes. The SURGE proposes that pain and stress in response to surgery are appraised through learned patterns of responses and what these responses will accomplish, response outcome expectancies. Negative response outcome expectancies sustain the activation of the physiological stress response and increase the risk of CPSP through pathophysiological mechanisms such as central sensitization, cortisol dysfunction, impairment of corticolimbic connectivity and inflammatory induced sickness behavior.

### Hypnosis

Hypnosis is a non-pharmacotherapeutic technique that holds promise as a harmless and effective pre-operative intervention to alleviate acute post-surgical pain and fatigue (31, 32). Clinical research with at least 20 different surgical populations has indicated that hypnosis can reduce the need for medication, reduce post-surgical symptoms, and enhance recovery (32). Furthermore, meta-analyses (33), narrative reviews (34, 35), and randomized clinical studies (36–39) all support the potential clinical utility of hypnosis with surgical patients. A rigorous trial from the US demonstrated particularly promising results (40). In that trial, a brief hypnosis session before breast cancer surgery had an impact on the patients’ need for pain medications and the level of postoperative pain, nausea, exhaustion, and discomfort, with moderate to large effect sizes. However, a French study, following a similar protocol but with some deviations from the American study, was not able to reproduce the effects (41). As such, there is a need for rigorous replication studies in different contexts. Moreover, no studies have so far investigated long-term effects of preoperative hypnosis, and whether it could in fact contribute to prevent acute post-surgical pain from turning chronic.

### Acceptance and Commitment Therapy (ACT)

ACT is an evidence-based treatment for chronic pain (42, 43) and is also a good fit for treating chronic fatigue (44). ACT is well suited to cancer populations as it offers a model of healthy adaptation to difficult circumstances and has shown promise in the treatment of opioid misuse (45). The goal of ACT is to increase psychological flexibility by reducing the influence from cognition and language when it produces an inability to persist or change in the service of long term valued goals. Psychological flexibility has been defined as the process of being in the present moment while persisting or changing behavior in accordance with chosen values (46). There have been several calls to expand the scope of ACT interventions to the treatment of CPSP (47). In a recent initiative, ACT was applied as treatment for CPSP in The Toronto General Hospital, with preliminary promising results. Those receiving ACT demonstrated greater reductions in opioid use and pain interference, as well as reductions in depressed mood, compared to those who received treatment as usual (48, 49). In addition to being an evidence-based treatment for pain and fatigue, ACT is also among the very few psychological treatments with documented effects on return-to-work (50).

Moreover, in a recent Norwegian study, a small-scale ACT follow-up intervention consisting of 1-6 phone calls, eased patients transition into the workplace (51). Even though both CPSP and chronic fatigue are common consequences of breast cancer surgery (7, 52), with disabling effects on recovery outcomes and quality of life, hardly any studies have looked at preventive interventions from a psychological perspective. As psychological risk factors are substantial, and the pathophysiology of both conditions largely unknown, interventions focusing on the psychological factors are largely needed.

## Methods/Design

### Study design

The PREVENT trial is designed as a randomized controlled, surgeon and statistician blinded superiority trial with two parallel groups and a primary end point of self-reported pain and fatigue 3 months after surgery. Randomization will be performed as block randomization with a 1:1 ratio. Participants are randomized to an intervention group including pre-operative hypnosis and post-operative web-based ACT (iACT), or a control group including pre-operative mindfulness and post-operative treatment as usual (TAU). We will thus be able to assess the short-term effectiveness of the pre-operative hypnosis alone, and the long-term effects in combination with the post-operative iACT. The aims are pursued with mixed methods, including an effect evaluation and a process evaluation.

### Study setting

The PREVENT trial is a single center trial placed in Oslo, Norway, in a large university hospital (Oslo University Hospital, Aker). In the Department of Breast and Endocrine Surgery, more than 400 breast cancer surgeries are performed every year.

### Aims and objectives

The overall aim of the study is to develop and test an easily implementable strategy of preventing chronic pain and fatigue after breast cancer surgery. The intervention strategy involves a pre-operative hypnosis session and a post-operative, internet-based Acceptance and Commitment Therapy (iACT). Together they form an intervention strategy with both a preventive and a rehabilitative focus. The PREVENT trial is designed to answer the following questions:

### Primary objectives

1. Will pre-operative hypnosis + iACT lead to less pain and fatigue 3 months after surgery compared to an attentional control?
2. Is hypnosis more effective than an attentional control in alleviating side effects from surgery? A replication study.

### Secondary objectives

The following research questions are exploratory but will be guided by previous findings and our theoretical model targeting secondary effects of treatment. Compared with our attentional control:

1. Will hypnosis impact levels of high sensitive c-reactive protein (hs-CRP) 3-4 weeks after surgery?
2. Will the potential effects of hypnosis in alleviating side effects sustain 3-4 weeks after surgery?
3. Will hypnosis in combination with iACT result in fewer prescriptions for pain and psychotropic medication in the 12 months after surgery?
4. Will hypnosis combined with iACT be favorable in a cost-utility analysis and result in fewer days of sick leave the following year after surgery?
5. Will hypnosis + iACT lead to increased psychological flexibility at 1-year follow-up?

In addition to secondary effects of treatment, we have the following research question exploring potential mechanistic pathways of pain and/or fatigue after surgery:

6. Does long-term stress (hair cortisol concentration), low grade inflammation (hs-CRP) and/or immunological reactivity (TruCulture supernatant) independently or in combination explain the development of chronic pain and/or fatigue after surgery?

Further, the following research questions explore the study cohort in terms of modifiable risk factors and novel predictors of pain and fatigue after surgery: Do biopsychosocial factors predict acute and sub-acute pain in our control group?

7. Do biopsychosocial factors predict acute and sub-acute pain in our control group? More precisely, do surgical procedure, presurgical pain, anxiety, depression, negative response outcome expectancies and low social perceived support predict pain intensity and pain unpleasantness immediately after, and 4 weeks after surgery while controlling for age?
8. Are medical, psychological, or social variables at baseline the strongest predictors of CPSP and chronic fatigue 3 and 12 months after surgery?
9. What is the prevalence of surgical fear? Associations with biological and psychological variables and development of a risk profile.

Finally, the following research questions will be pursued in a qualitative process evaluation:

10. What types of lived narratives are associated with post-surgical pain and fatigue, and what types of narratives are associated with high quality of life and return to life as normal?
11. Are these narratives changed by the interventions, and if so, how?
12. How does pain catastrophizing fit into a person’s overall narrative, and (how) is it changed by the interventions?
13. How do different forms of narratives contribute to developing or sustaining post-surgical pain or fatigue?

### Hypotheses of efficacy

1. Compared to attentional control, hypnosis + iACT will be superior in preventing chronic post-surgical pain and fatigue 3 months after surgery.
2. Compared to attentional control, hypnosis will be superior in alleviating side effects from surgery.

### Measurements

Patient reported outcome measures (PROM) will be collected through a battery of 10 validated questionnaires, all relevant to the various aspects of breast cancer surgery and the post-surgical trajectories. It will be given to the participating women at baseline and at 3- and 12-months follow-ups. To secure high compliance, participants will receive 4 weekly reminders to fill out the questionnaires. The reminders will be sent both as a text message and through email. Our user representative reviewed an earlier version of the questionnaire package and provided input before it was finalized. Estimated time of completion is 15-20 minutes.

### Primary outcome measures

#### Chronic post-surgical pain

CPSP will be measured through the Numeric Rating Scale (NRS) for pain intensity. The NRS is a discontinuous, self-report measure. It consists of a single item, in which the respondent is asked to rate the intensity of pain. This rate is done on an 11-point scale ranging from 0 (no pain) to 10 (worst pain imaginable). In the context of pain, the NRS has been proven to have good reliability and validity (53, 54). It is a valid measure of pain intensity (0–10) that is less influenced by present mood state (55), and has been widely used to assess post-surgical pain in previous studies of women with breast cancer (3, 7, 8, 56). In the PREVENT trial, participants will be asked to rate their pain intensity in and around the surgical site and referred area (breast, axilla and arm), in line with the definition of CPSP (16). They will be given a total of 4 NRS questions about pain intensity in the breast, axilla, arm, or other parts of the surgical area. In the main analyses of efficacy, CPSP will be analyzed as a continuous variable where the worst pain at any site indicates the highest numerical rating scale value (0–10) reported by the patient. This way of operationalizing CPSP after breast cancer surgery is in line with previous studies in the field (26, 57).

#### Chronic post-surgical fatigue

Chronic fatigue will be measured through the 13-item Functional Assessment of Chronic Illness Therapy-Fatigue subscale (FACIT-F). The FACIT-F is a unidimensional self-report scale meant to assess fatigue and its impact on daily life. Consisting of 13 items, the scale asks the respondents to rate their level of symptom intensity on a 5-point scale ranging from 0 (not at all) to 4 (very much) as it applies to the past 7 days (58, 59). FACIT-F is a widely used measure of fatigue in breast cancer trials. It has demonstrated excellent internal consistency, high validity, and sensitivity to pick up change in patients with breast cancer (58). The total score ranges from 0-52, with higher scores indicating less fatigue. A score of less than 30 indicates severe fatigue. In the PREVENT trial, the main analysis will involve a comparison of the total score (0–52) 3 months after surgery between the intervention and control group.

### Secondary outcome measures

#### Side effects from surgery

A range of side effects from surgery will be assessed using the 100mm Visual Analogue Scales (VAS) after surgery, upon discharge, in line with the trial that we are replicating (40, 60). Both pain intensity and pain unpleasantness will be assessed to capture its sensory and affective dimensions. The VAS is a continuous, self-report measure comprised of either a horizontal or vertical line measured at exactly 100 mm. The line is anchored by verbal descriptors, and the respondent is asked to rate the intensity of their symptom by making a mark on the line. The distance between the end of the line, representing absence of the symptom, and the mark will be measured and a score ranging from 0 to 100 will be recorded. Similar to the NRS, the VAS can be used on a variety of symptoms and will here be used to assess pain intensity, pain unpleasantness, fatigue, nausea, discomfort and emotional upset (53).

#### The European Organization for Research and Treatment-QOL questionnaire for breast cancer specific module (EORTC QLQ-BR23)

The EORTC QLQ-BR23 is a health-related quality of life measure specifically designed for women with breast cancer. It consists of 23 items, each asking the respondent to indicate the extent to which they have experienced certain symptoms or problems on a 4-point scale ranging from 1 (not at all) to 4 (very much) (61). It has been shown to have high reliability and clinical and cross-cultural validity as a measure of quality of life in patients with breast cancer (61). This measure was specifically included upon recommendation from our user representative, and is measured at baseline, 3- and 12 months follow-up.

#### Hospital Anxiety and Depression Scale (HADS)

The HADS scale is a measure of anxiety and depression specifically designed for patients with physical illness by excluding somatic items (62). It includes 14 items that are equally divided into two subscales: depression and anxiety. Respondents are asked to respond on a 4-point scale ranging from 0 to 3. Consequently, each subscale has a possible score of 0-21, and a cutoff score of 8 or more on both subscales has previously been reported to give an optimal balance between sensitivity and specificity (∼0.80 for both subscales) according to DSM-III, DSM-IV, ICD-8, and ICD-9.29 (63). However, in a recent meta-analysis a HADS-D cut-off value of seven or higher were found to maximise combined sensitivity and specificity (64). The cut-off will therefore be 7 in the current study. HADS has demonstrated high reliability and validity across studies (63), and is in the current study included at baseline, 3- and 12-months follow-up.

#### Patient Global Impression of Change (PGIC)

The Patient Global Impression of Change aims to measure the patient’s subjective experience of change in function, symptom and quality of life over time, and are meant to reflect the patient’s belief about the efficacy of the ongoing intervention (i.e., “feeling better” or “feeling worse”) (65). The PGIC is constructed of a 7-point scale where patient rate their change as “very much improved”, “much improved”, “minimally improved”, “no change”, “minimally worsened”, “much worsened” and “very much worsened” and is measured at 3- and 12 months follow-up. In studies of pain populations, it has been demonstrated that the “much improved” and “very much improved” ratings indicate moderately important and substantial improvement (66), and PGIC has also shown clinical relevance in daily clinical practice (67).

#### Acceptance and Action Questionnaire (AAQ-II)

The Acceptance and Action Questionnaire is designed to measure psychological flexibility. It consists of 7 items measuring levels of acceptance and avoidance of unpleasant thoughts and feelings (68). The items are scored a 7-point scale ranging from 1 (never true) to 7 (always true). The AAQ-II has showed good reliability and validity across a range of linguistic and clinical populations, including Norwegian samples and among patients with breast cancer (69, 70), and will be measured at baseline, 3- and 12-months follow-up.

#### Perceived social support

Perceived social support is operationalized based on the conceptual framework on social relations as described by Due et al. (71) and as formulated as specific questions by Skovbjerg et al (72). The structure is meant to measure the emotional support and practical assistance expected to be available, by asking the following question “will any of the following people help or support you in everyday life, if you need it?” in regard to “partner”, “family”, “friends”, “colleagues”, or “neighbors”. For the current study, we decided to include a sixth option; “others”, to the list of possible sources of support. Patient rate the perceived social support of these relations respectively using a 6-point scale ranging from 1 (always) to 6 (never or have none) (72). Social support will be measured at baseline, 3- and 12-months follow-up.

#### Injustice Experience Questionnaire (IEQ)

The IEQ is a 12-item measure meant to assess the respondent’s experience of injustice (73). Injustice encompasses the degree of blame as well as the magnitude and irreparability of loss related to their health condition. The items therefore consist of thoughts and feelings related to injustice. Respondents are asked to rate their experience of injustice on a 5-point scale ranging from 0 (not at all) to 4 (all the time) (73). The scale has been translated and validated in Norwegian (74). In the PREVENT trial we slightly adjusted the wording in the scale instruction to make it fit patients diagnosed with breast cancer. Further, to minimize the burden of filling out long questionnaires in a vulnerable situation, we constructed a short version of the IEQ with 5 items. The 5-item version of the scale ranges from 0-20 and have a mean of 8.3 (SD 5.7). These numbers are based on data from 3170 patients with various chronic pain conditions. As a test of validity, the Pearson correlation between the 5-item version of the IEQ and the total score of the full version of PCS was tested: r=0.703 (p<.01). This is similar to the correlation between the full scales of each measure (r=0.721) and is as such an indication of scale validity. IEQ is measured at baseline, 3- and 12-months follow-up.

#### Bergen Insomnia Scale

The Bergen Insomnia Scale (BIS) will be used to assess insomnia symptoms. This questionnaire consists of six items, of which the first four pertain to sleep onset, maintenance, early morning wakening insomnia, and not feeling adequately rested after sleep, corresponding to the DSM-IV criterion for insomnia (75). The last two items assess level of daytime impairment due to sleepiness and satisfaction with sleep, corresponding to the DSM-IV criterion B (75). Each item is rated on average occurrence from 0–7 days per week, giving a possible total sum score from 0 to 42. The Bergen Insomnia Scale has shown high reliability and validity in both patient and community samples (76). Insomnia will be measured at baseline and 12 months follow-up.

### Registry data

Registry data on employment and benefit take-up and medical prescriptions will be obtained from the Norwegian Labor and Welfare Administration (NAV) and the Norwegian Prescription Database (NorPD), respectively. To be able to assess for the effects of the interventions, data from NorPD and NAV will be obtained from two years prior to inclusion in the trial until one year follow-up. The NorPD contains data about dispensed drugs in Norway, while NAV-data contains social security micro data for research. These data will be obtained retrospectively.

### Biomarker variables

#### Stress reactivity

The immune functional assay will consist in collecting whole blood samples using the standardized TruCulture system (Myriad RBM, Austin, Texas, USA) directly containing immunogenic stimuli. There are two tubes, one containing a control medium (Null tube) and the other lipopolysaccharides (LPS) at 100 ng/mL (from Escherichia coli O55:B5). Immune function is to be assessed by analyzing the supernatant fluid. TruCulture® reproducibly reveals the induced innate and adaptive immune response in whole blood after stimulation, by quantifying the release of soluble immune activation products (cytokines, chemokines, soluble receptors etc.) in the supernatant and by measuring the transcription level (mRNA) in the circulating blood (immune) cells (77). TruCulture samples will only be collected pre-surgery and investigated as a possible predictor of chronic pain and fatigue.

#### Low-grade systemic inflammation (hs-CRP)

The high sensitive CRP (hs-CRP) test accurately measures low levels of CRP to identify low but chronic levels of inflammation. It measures CRP in the range from 0.5 to 10 mg/L. Non-fasting serum samples will be drawn from all participants both pre- and post-surgery (3-4 weeks after surgery) and analyzed at the Department of Laboratory Medicine at Aker Hospital in Oslo, Norway.

#### Hair cortisol

For a few years now, a new and very different method to measure cortisol exposure in humans has been developed; the extraction of cortisol from human hair, with major advantages involving the non-invasive nature, the standardized sampling, and most importantly, the possibility to use hair as a retrospective biomarker of cortisol exposure (78). Hair grows approximately one centimeter per month, thus making it possible to show the average long-term activity of the HPA-axis, as well as compare hair segments/months with each other (e.g., before and after stressful events). Another advantage is that hair cortisol reflects the amount of free unbound cortisol, which is unaffected by oral contraceptives (79).

### Background variables and clinical characteristics

Besides from demographic characteristics such as age, education, occupational status, marital status and number of children, the following variables will be measured at baseline:

#### Clinical data

Clinical information prior to surgery will be collected by the study nurses during the inclusion appointment. This includes information on height, BMI, smoking, number of previous surgeries (in breast area, and other areas of the body), previous diagnoses of cancer or any chronic pain conditions. Patient charts will be used to collect data on surgical procedures and potential complications during/after surgery. This will include information about the duration of the surgery, tumor-related variables, severity of acute post-surgical pain, and other relevant intra-operative variables, such as type of surgical procedure (i.e., breast conserving surgery, mastectomy, breast reconstruction, sentinel node biopsy, axillary lymph node dissection, nerve blockage and surgical drain), and if there have been any surgical complications. Further, it is noted if patients have received neo-adjuvant treatment, and if their surgery is performed as a day case or requires post-surgical hospital admittance. Medications used during/after surgery is also recorded. This involves intraoperative use of analgesics for pain control and of sedatives, as well as postoperative use of medications for pain control. These are in line with the outcomes reported in the trial we are replicating (40).

#### Widespread pain questionnaire

As a measure of pre-existing pain we used a modified version of the Fibromyalgia Survey Diagnostic-2016 criteria (FSD-2016 criteria) (80) translated and validated for Norwegian participants (81). The modified questionnaire asks about pain in the four quadrants of the body (left upper, left lower, right upper, right lower), if this has persisted for more than 3 months and if it is bothersome, it also covers head and visceral pain to exclude this from the picture, giving a short and pragmatic index of chronic widespread pain.

#### The Pain Catastrophizing Scale (PCS)

The PCS is a self-report measure of catastrophizing. The scale assesses elements of rumination, magnification, and helplessness in response to pain (82). Responders rate items on a 4-point Likert scale ranging from 0 (not at all) to 4 (all the time). To decrease responder burden, this study uses the short version of the scale (PCS-4). PCS-4 has been validated in surgical (83) and chronic pain populations (84).

#### Pain expectations

A single item is used to assess how much pain the women expect to have after surgery. A single-question approach has previously shown high predictive validity after cesarean delivery (85). The participant will be asked to rate the intensity of pain she expects to have after surgery on an 11-point scale ranging from 0 (no pain) to10 (worst pain imaginable).

#### Return to work expectations (RTW)

Return-to-work expectations (RTW-expectations) is assessed by asking participants to respond to the following statement: “I expect to return to work within the next few weeks”. Responses are given on a five-point Likert scale ranging from “strongly agree” to strongly disagree”. In line with previous studies, we will trichotomize the variable in to positive, uncertain, or negative RTW-expectations (86, 87). This single item measure of RTW-expectations has been demonstrated to be a strong predictor of non-RTW in populations with similarities to the current, such as people on sick leave due to chronic low back pain (88), and people struggling with work participation due to common mental disorders (86).

#### Job satisfaction

Job satisfaction will be assessed with two single items used in several previous trials (e.g., 89). The first item asks participants how satisfied they are with their overall job situation, where they indicate their response on a Likert scale from “very satisfied” to “not satisfied at all” or “not currently working”. The second item asks what job they would choose if they could choose again, with the alternatives “would choose my current job”, “would choose not to work at all” or “would choose a different job than the one I have now”. Job satisfaction is measured at baseline, 3- and 12-months follow-up.

#### The Surgical Fear Questionnaire (SFQ)

The SFQ is a reliable and valid self-report instrument designed to assess fear of surgery, and it consists of 8 items which is scored on an 11-point scale ranging from 0 (not at all afraid) to 10 (very afraid) (90). The items target different fears concerning surgery, namely fear of operation, anaesthesia, postoperative pain, side effects, health deterioration, failed operation, incomplete recovery, and long duration of rehabilitation. The structure of the SFQ can best be described by a two-factor model, in which two subscales can be distinguished: fear of immediate consequences of surgery and fear of the long-term consequences. We translated the scale to Norwegian through a forward-back-translation procedure.

#### Life Orientation Test – Revised (LOT-R)

The LOT-R is a 10 item self-report test meant to assess dispositional optimism. The respondents are asked to rate their agreement to various statements on a 5-point scale ranging from 0 (strongly disagree) to 4 (strongly agree). Three of the items assess optimism, three assess pessimism, and the remaining four are filler items that are not meant to be included in the scoring (91). A validated Norwegian version of the scale will be used in the current study (92).

**Fig 1.**
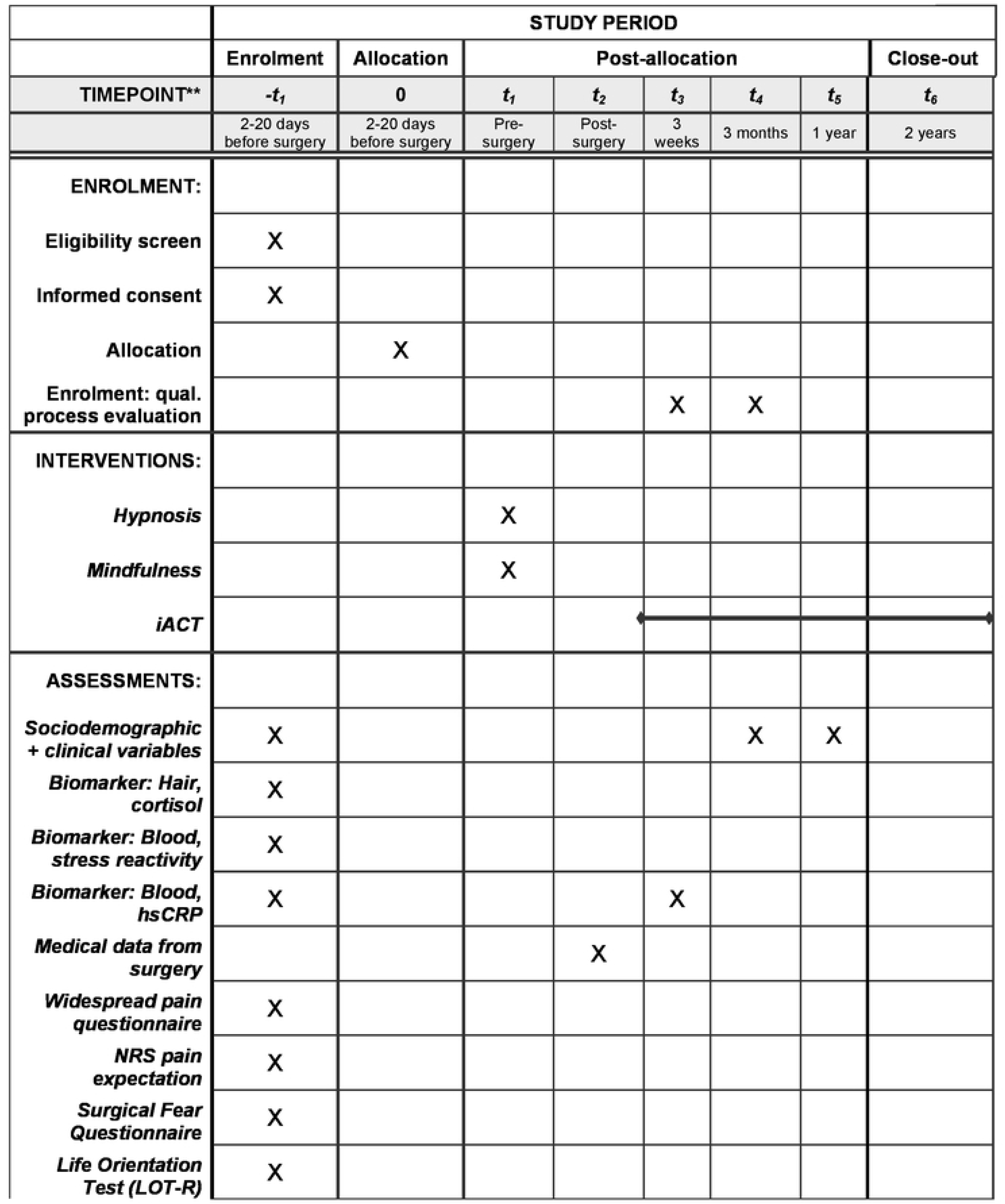
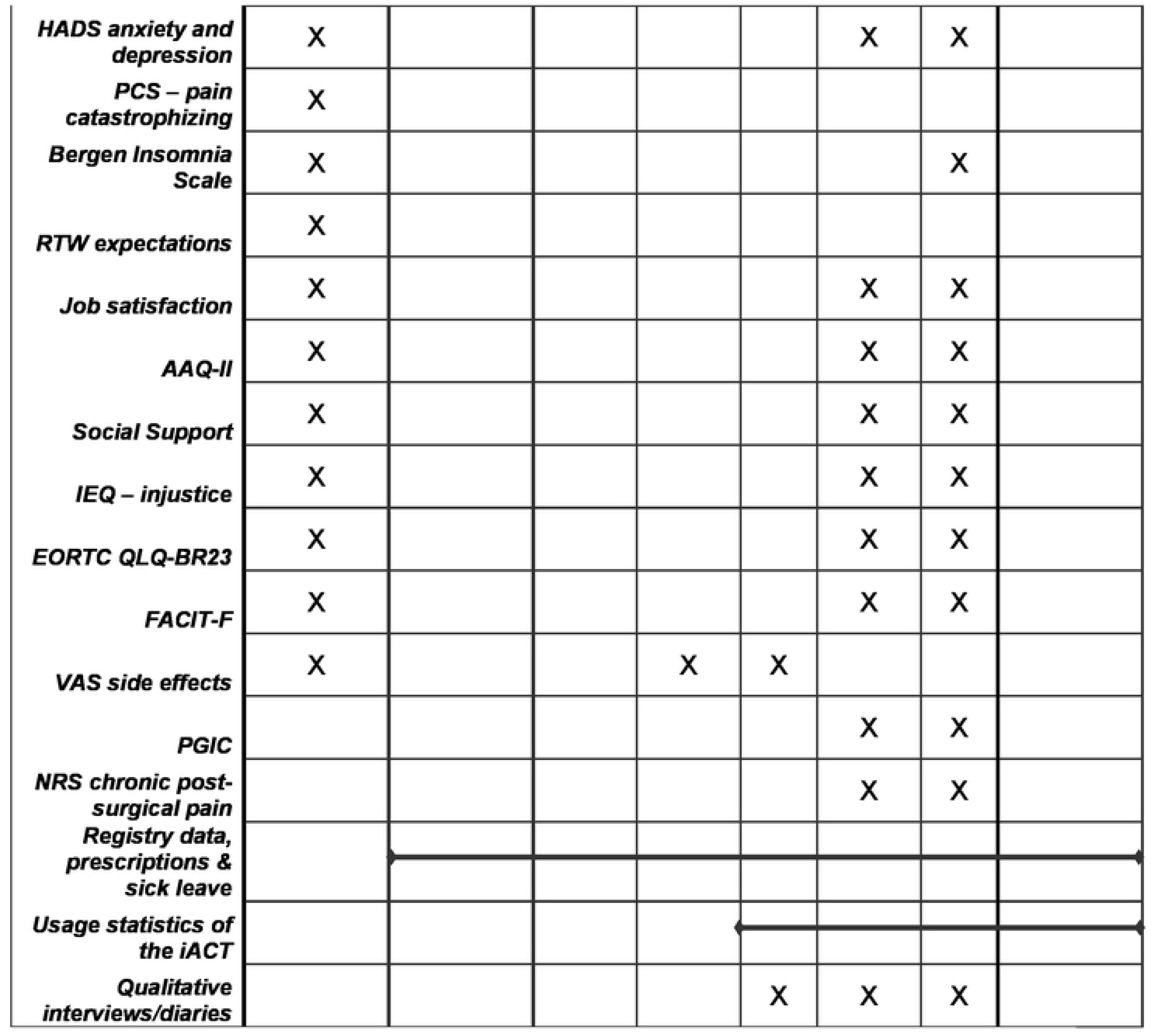
Schedule of enrolment, interventions, and assessments.

**Figure 2.**
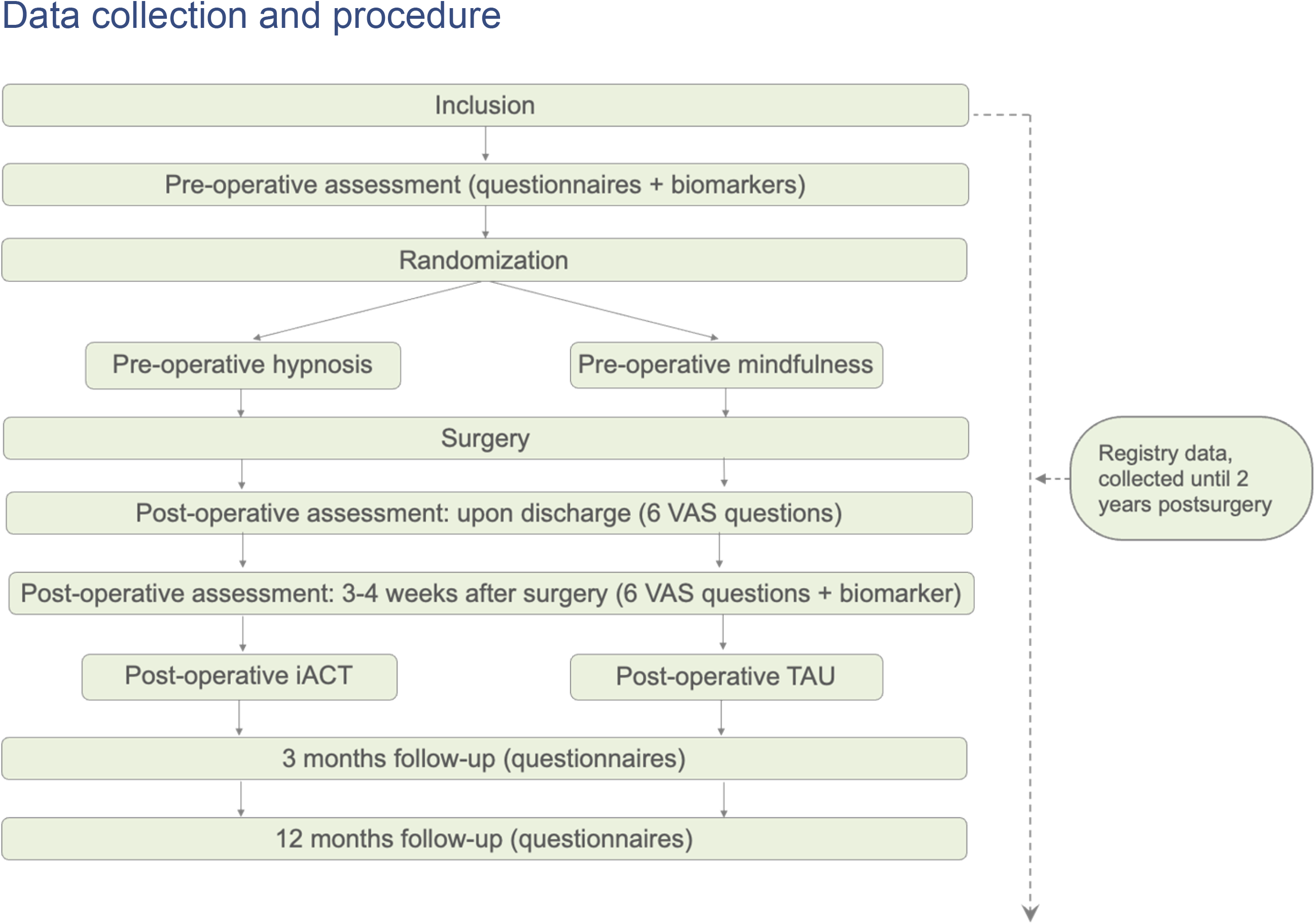
Flow chart of the study logistics and data collection.

### Before surgery

Patients scheduled for breast cancer surgery at the Department of Breast and Endocrine Surgery, Oslo University Hospital, Aker, receive oral and written information about the PREVENT trial from the doctors or nurses responsible for their treatment. This usually happens at their first regular consultation at the hospital, following diagnosis. If patients are interested in participating, they immediately or within the first following days and prior to surgery receive a consultation with a study nurse. The study nurse provides the patients with information about the study and go through the inclusion- and exclusion criteria. Patients who are eligible and want to participate sign the informed consent form, in which case the study nurse introduces the patients to the online data collection platform, VieDoc^TM^, which includes the questionnaires (PROMs) that must be filled out (once before surgery, 3- and 12 months after surgery). The questionnaires can be filled out at home using the patient’s own electronic devices (VieDocME^TM^). In VieDoc^TM^, patients are automatically randomized (ratio 1:1) to one of two groups: Intervention (Pre-surgical clinical hypnosis and post-surgical iACT), or Control (pre-surgical mindfulness and post-surgical TAU). Further at this point, the study nurse collects hair and blood samples from the patients. At the day of surgery, patients in the intervention group are met by a trained specialist in clinical hypnosis, who provides the 20-minute hypnosis session. Patients in the control group are met by a study nurse, who plays a pre-recorded mindfulness session (13 minutes) delivered on an iPad with headphones. All treatments (hypnosis, mindfulness and iACT) take place in an undisturbed designated room at the hospital ward. The patients will receive the hypnosis/mindfulness sitting in a chair or lying in a bed, depending in their own preference. Immediately after the treatment, patients are taken to the operating room receiving their anesthetic and surgical procedures.

### Immediately after surgery

Post-surgery, after waking up and before discharge, the patient fills out six VAS questions on pain intensity, pain bothersomeness, fatigue, nausea, discomfort, and emotional distress. This is filled out with pen and paper, and the questions are identical to the questions used in the trial we are replicating (40).

### 3-4 weeks to 12 months after surgery

3-4 weeks after surgery, as a part of a regular outpatient consultation at the hospital, a study nurse repeats blood samples and the six VAS questions. After the questions (PROMs) are answered, patients in the hypnosis + iACT group are introduced to the post-operative intervention (iACT) and instructed on how to use the iACT platform. Patients in this group can access the platform from home during the complete one year follow up period. All patients are reminded via e-mail to use VieDocME^TM^ to answer the follow up questionnaires 3- and 12-months after surgery. The 3-month timepoint where patients provide PROMs was chosen as this aligns with the time when CPSP is diagnosed. The 12-month timepoint of PROMs collection was chosen to align with previous studies on long-term follow-up.

### Allocation

The allocation sequence is computed-generated by VieDoc^TM^, where patients are randomized (ratio 1:1) to one of two groups: Intervention (Pre-surgical clinical hypnosis and post-surgical iACT), or Control (pre-surgical mindfulness and post-surgical TAU). Block sizes will vary between 4, 6 and 8. The study nurses in charge of enrollment do not have information about block sizes. As soon as a patient is enrolled and has completed the baseline questionnaire (PROMs), the allocation of that patient is generated automatically in VieDoc to the study nurse who then assign the patient to the intervention and inform about the procedure.

### Blinding

Blinding of participants is not possible due to the character of the interventions. However, the following personnel will be blinded: The surgeons performing the surgery, the anesthesiologists and nurses, and the trial statistician, who is also the outcome assessor. The person who accompanies the patient to the surgical ward is instructed not to reveal the allocation to the health care personnel at the surgical ward. The participant is also instructed not to reveal which pre-operative intervention they received.

### Drop-out and strategies for PROM data retention

Participants who no longer wish to participate in the study can inform the research group of their decision by notifying the study nurse, the trial coordinator, or the PI. The trial coordinator or the study nurse will in any case contact participants who drop out by phone and ask if they are willing to report the reason for withdrawal. If reasons are provided, these will be registered on a dedicated drop-out form. They will then be presented with the following options: First, they will get the option of discontinuing the interventions only but keep receiving the questionnaire follow-ups. If they do not wish to hear from us again, they will be registered as dropouts from the trial, but we will ask for permission to use already collected data, as well as follow them up with registry data. Lastly, if they wish to be completely excluded from the trial and have all their data deleted, we will adhere to that. Through this differentiated strategy we hope to retain as much PROMs data as possible from participants who drop out from the trial. Participants who drop out of the study will still be included in the intention-to-treat (ITT) analyses, unless they required all their data deleted.

### Data management

Data from all sources will be entered and stored on a secure server. The University has an approved system for that which we will use (TSD). When recruitment and data collection is concluded, data will be extracted from VieDoc and imported in to TSD where a designated data manager will clean the data and prepare them for analyses. A detailed data management plan is available for download (93).

### Interventions

#### Pre-operative hypnosis

The pre-operative hypnosis protocol is identical to the study we are replicating (40). The scripted session includes a relaxation-based induction, suggestions for pleasant visual imagery, suggestions to experience relaxation and peace, and specific symptom-focused suggestions. The intervention has been thoroughly translated and piloted in a previous study that showed both feasibility and acceptability of hypnosis in this context (94). Trained psychologists and last-year psychology students will provide the hypnosis in a dedicated room in the hospital. Patients will be encouraged to use self-hypnosis on their own following the treatment session. The hypnosis lasts for 15-20 minutes and is provided within 2 hours prior to surgery. The patients will be invited to either sit in a comfortable chair or lay down in a bed during the hypnosis. After the session, the therapist or the study nurse follows the patient to the surgical ward.

#### Active control condition

Patients randomized to the control group will receive a mindfulness before surgery. The mindfulness session will be delivered through a pre-recorded audio file on a mobile phone with headphones. The audio file guides listeners through a 13-minute mindfulness session where they are invited to sit/lay down and pay attention to their surroundings, before being invited to notice their breath. The subsequent invitation is to count inhales and exhales until you reach 10, and then starting over again. The counting is prompted throughout the rest of the session until 13 minutes have passed. Recorded mindfulness meditation have shown significant effects on chronic pain and fatigue in breast cancer patients (95). However, the referred studies all entail numerous sessions of mindfulness with some degree of therapist involvement. We therefore believe a single session of mindfulness delivered through an audio file, constitute an active and ethically sound control condition. The patients will be invited to either sit in a comfortable chair or lay down in a bed during the mindfulness session. After the session, a therapist or the study nurse follows the patient to the surgical ward.

#### Comparison of interventions

*Pre-surgery intervention versus active control:* Hypnosis and mindfulness are commonly regarded as supplements or integrative parts of an ACT intervention (43). They both start by focusing the patient’s attention in similar ways but proceed to utilize that focused state into different means. The pre-operative hypnosis here is used with a specific purpose and intent of lessening pain and promote coping. The pre-operative one-session mindfulness has a specific purpose and intent to focus on one thing like the breath, sounds or the body. Thus, both techniques guide the patients experience and use suggestion, but the end goals are fundamentally different.

### Criteria for discontinuing allocated interventions

Before the hypnosis and mindfulness session, patients are informed that they at any point, and without stating any reason, can disrupt the session. They are informed about the harmless nature of the interventions, with few or none reported side effects, but they are also informed that if they should experience any discomfort or other adverse events and wish to terminate the session, the treatment will be immediately terminated. The occurrence of potential adverse events will be registered by the therapist administering the treatments, as well as registered through PROMs (PGIC) at 3- and 12-months follow-up.

### Post-surgical intervention

The post-surgical intervention is comprised of a web-based ACT intervention (iACT) with the cognitive activation theory of stress as theoretical vantage points (29). The intervention consists of videos in combination with a work booklet targeting three main processes derived from the ACT hexaflex model (46), and delivered in a web based interface where a specific order of viewing is suggested, but not enforced. In the suggested order, the intervention has several videos that is built to involve the patient in a broadening of their perspective of what can constitute maintaining and debilitating factors when experiencing chronic pain and/or fatigue. These initial videos involve a biopsychosocial explanation of pain and fatigue, alongside an introduction to mindfulness, values-based living, and stress reduction through cognitive behavioral techniques.

The post-surgical intervention also involves the use of a hand-out booklet that serves as an addition to numerous videos presented to the participants. The videos are sorted in three categories aimed to communicate the six processes in a commonsensical way. The three categories are referred to as the tri-flex in the Focused ACT model (96), and consists of Aware (present-moment awareness and self-as-context), Open (Accept and defusion) and Engaged (values and committed action). Within the booklet there are written tasks corresponding to different videos and scenarios shown in the online intervention to approximate previous trials showing efficacy on chronic pain from a similar intervention (97).

Following the call of Bewick and colleagues (98) for a more standardized reporting of intervention content, iACT can be described as a web-based intervention targeting women undergoing breast cancer surgery. It is delivered in secondary care, free of charge, but currently only for this research project. Participants choose how much time to spend on the intervention and can use it for as long as they like (up to a year). Each video lasts approximately between 5 and 25 minutes. It has a strong theoretical foundation, based on the CATS-model as well as the ACT model. The interventions are not tailored and includes no counselor involvement. In terms of information architecture (99), the intervention has a matrix design (participants can navigate freely), with topically organized content. After the last PROMs data collection, at 1 year follow-up, participants in the control group will be offered access to the iACT platform.

### Adherence to intervention protocols

Regular supervision will be conducted with the therapists conducting the pre-operative hypnosis to secure compliance to the protocol, and every second hypnosis session is audio recorded for quality assurance. We plan to review a random selection of audio recordings, at least 20% in total, to assess adherence to the hypnosis script. To assess adherence to the post-operative intervention, we will have statistics on each participant’s program use, measured by what videos/audio files they have seen/listened to, and how many times they have accessed the various content.

### Concomitant care

All participants in the trial will receive care as usual in addition to the interventions that they receive in the PREVENT trial. This could involve medical treatment such as chemotherapy, radiation, endocrine treatment etc., as well as psychological treatment and support. There are no treatment restrictions attached to participation in the PREVENT trial. Concomitant treatment will, however, be registered.

### Participants

The study will recruit and randomize 200 participants amongst patients from the Department of Breast and Endocrine surgery at Oslo University hospital (see sample size calculations for justification).

### Inclusion criteria

Patients must fulfill the following criteria to be eligible for inclusion in the study:

- Women diagnosed with breast cancer and scheduled for surgery
- Be able to provide informed consent
- Over the age of 18

### Exclusion Criteria

- Insufficient Norwegian speaking or writing skills to participate in the interventions and fill out questionnaires
- Over the age of 70
- Cognitive and psychiatric impairment or other serious malignancies

### Sample size and power

Sample size is estimated for both primary outcomes. Previous trials with similar interventions have demonstrated moderate/large effect sizes on pain and fatigue (60, 100). However, compared to these studies, we expect more heterogeneity in our data as our study population will undergo several medical treatments during the trial (e.g., radiotherapy or chemotherapy after surgery). We thus expect a minimum difference that is clinically relevant to be 1 point on NRS (0–10) and 3 points (which corresponds to patients reporting being “slightly better”) on FACIT-F (0–52). Based on previous studies, it is expected that the control group scores 5 (SD 2.4) on NRS at 3 months follow-up (101), and 18 (SD 7) on FACIT-F (60). With a power of 0.80 and a two-tailed α of .05, the number needed in each group to detect the difference in pain is n=92, and in fatigue n=87. To account for attrition and loss to follow-up, we plan to include 100 patients in each group.

### Statistical analyses

Effect analyses of primary and secondary outcomes will be performed according to both the intention-to-treat principle (the primary analysis) and per protocol. The efficacy analysis of the primary outcomes will involve a comparison of mean pain and fatigue levels at 3 months follow-up by use of independent sample t-test. This will constitute the main analysis of the primary outcomes. Additionally, they will be analyzed using linear mixed models (also referred to as multilevel models) at the 3 different time points (post-surgery, 3- and 12-months follow-up). Multilevel modelling is a flexible statistical approach that can handle non-balanced data with missing entries and repeated observations (102).

### Sub-group analyses

Any significant main effects between the intervention and control group may allow for subgroup analyses. The following pre-defined subgroups will be investigated: Surgical procedure (breast conserving vs mastectomy), axillary lymph node dissection (yes/no), pre-existing pain condition (yes/no) and clinically relevant anxiety. All these variables have been shown to predict CPSP and other late effects from breast cancer surgery and could as such act as moderators of any treatment effect.

### Handling of missing data

For the analyses of secondary outcomes, we plan to apply mixed effects models by use of Maximum Likelihood Estimation (MLE) which is a recommended approach to handle complex structures of missing data. In MLE, there will be a robust adjustment for missing observations, accounting for complex structures of missing data (103). Due to the considerable number of secondary outcome measures in the PREVENT trial, alpha inflation is a concern, and these analyses will as such be interpreted with caution.

### Implementation

The project is administered from the Dept of Psychology, University of Oslo, but data will be collected, stored, and analyzed at Oslo University Hospital through VieDoc which is an established and secure system for data collection, storing and management approved by the hospital. The trial runs at the hospital in the Department of Breast and Endocrine surgery. Two study nurses are employed and currently work in the out-patient department and on the ward with patient recruitment, follow-up, and biomarker assessments. A research assistant is responsible for practical aspects of the trial as well as administering the web-based platform where the iACT is delivered. An experienced psychologist with expertise in hypnosis is responsible for delivering the pre-surgery hypnosis and supervising the other therapists (psychologists/last-year psychology students). They have all gone through didactic and practical training in the specific hypnosis intervention used in the trial, as well as completed supervised practice as part of their training. The iACT was developed under the leadership of the co-PI of the study (Jacobsen), with input from our national and international partners (Linton & Flink), and user representatives. The user representatives have also provided valuable feedback upon completion of the post-operative intervention that has led to several adjustments and modifications.

To disseminate the results to patients, clinicians, policy makers and the public, we are planning to co-host a seminar with the Norwegian Cancer Society for stakeholders and collaborators to present results from the trial. The Cancer Society will also communicate the research findings through their channels, which are well organized and far reaching.

### Process evaluation

#### Participants

Participants in the RCT will also be invited to contribute to the qualitative process evaluation. We expect to include 6-16 participants in the process evaluation, with an equal number of participants from each study arm. With qualitative methodology, the general rule is that quality is more important than quantity – that is, having many participants is less important than ensuring high quality of the data and the analysis (104). Furthermore, the final number of participants can rarely be decided upon in advance, but rather, the decision to stop inclusion depends on the quality of the data collected and the needs of the analysis. Patients participating in the process evaluation will sign a separate informed consent relating to participation in this ancillary study.

Participants will be recruited in two waves. In the first wave of recruitment, participants will be purposively selected on the basis of maximum variation on expectedly important parameters (105), such as age, type of operation, and whether the cancer is recurrent or new. In the second wave, participants will be selected based on their scores on the pre-operation questionnaire in the quantitative part of the study, specifically their scores on pain catastrophizing. We will purposefully select some participants who score high on pain catastrophizing and some who score low, with the purpose of unpacking this predictor qualitatively and understanding how it may be changed by the interventions.

#### Form of data collection

Data will be collected through individual, semi-structured interviews and a diary. The interviews may be conducted face-to-face, over the telephone, or via Zoom, and the participants will be asked about her thoughts and feelings about the past, present, and future in relation to the cancer diagnosis, the treatment, and quality of life. At the first interview, around 4 weeks after surgery, participants will be given a diary to record their thoughts and feelings pertaining to the illness, treatment, symptoms, their situation, and the future. The diary will be collected before the next interview, which will take place some months later.

#### Analyses

Data will be transcribed by the researcher and analyzed with two separate analyses: (socio)narrative analysis (106, 107) and Grounded Theory (108). The different analyses are expected to result in different, complementary perspectives on the development of CPSP and fatigue and how the intervention strategy influences these processes. In reporting from the process evaluation, we will follow the journal reporting standards for qualitative research (109).

### Confidentiality

PROMs will be completed electronically using the ViedocMe functionality available in Viedoc. Study staff will create a ViedocMe account for each participant in the participant’s Clinic View in Viedoc and provide a unique log-in profile (username, pin-code, and ViedocMe web-address) for each participant.

### Data protection and management

All data will be stored on a secure server at University of Oslo (TSD). This is a platform for collecting, storing, analyzing, and sharing sensitive data in compliance with the Norwegian privacy regulation. A detailed plan for data management has been developed in collaboration with both Oslo University and the University of Oslo that shares the responsibility for data management in this study. Only the PI, the co-PI and the data manager will have access to all the collected data. Other researchers will be granted limited access according to their need of data for their research. TSD has a system in place for differentiated access.

### Dissemination policy

The results will be disseminated to the international academic community through publications in peer-reviewed scientific journals, adhering strictly to the Vancouver Protocol. In collaboration with our user representative in the research team, we will strategize a plan to communicate the findings more broadly. This could involve seminars, workshops and lectures for health care providers and other stakeholders, articles in the broader news media, and feature articles in specific user magazines or websites and conferences for health workers, health authorities, policy makers, and researchers. Our group follows an active media policy, providing that results have been published in scientific journals, with presence on several social media platforms. We have developed a website dedicated to the PREVENT trial where we will communicate information about the project’s key findings and conclusions. Finally, we plan to share anonymous README files as well the statistical codes for the main analysis in an open repository (e.g., Open Science Framework) to ensure transparency and reproducibility.

## Discussion

Breast cancer is the most common form of cancer among women (1), and a majority of women report persistent psychological and physiological symptoms after breast cancer surgery (3–5). Chronic post-surgical pain and fatigue are the most common sequelas (6, 13). This is a both a major clinical problem, with prevalence rates ranging up to 60% (6), and a socioeconomic burden on the health care system (110). Studies on preventive interventions are scarce, and those that have been carried out have not been sufficiently replicated.

This study will to our knowledge be the first study to investigate the combined effects of a pre-operative hypnosis intervention and a web-based post-operative ACT intervention in women about to undergo breast cancer surgery. It will also be among the very few attempts worldwide to prevent the incidence of pain and fatigue after breast cancer surgery applying a psychological approach. This study design has several strengths. The outcome measures are valid and reliable, involving both self-report measures, objective registry data, and biomarker data, as well as a qualitative process evaluation sub study, and the study will contribute to the need of rigorous clinical trials within this field of research. The project results will be highly relevant to all health care personnel involved in breast cancer care, but also to the larger community of clinicians and researchers within the field of chronic pain and fatigue.

### Trial Status

The trial started recruitment in October 2020 and is still collecting data for the primary endpoint.

## Data Availability

Data will available from the NSD Archive (contact via https://www.nsd.no/en/find-data) for researchers who meet the criteria for access to confidential data.

## Abbreviations

ACT: Acceptance and Commitment Therapy
CPSP: Chronic Post-Surgical Pain
EQ-5D: European Quality of Life – 5 Dimensions
QoL: Quality of Life
RCT: Randomized Controlled Trials
TAU: Treatment As Usual

## Declarations

## Acknowledgements

We would like to express our gratitude to all our collaborators who contributed with valuable input to the planning of the PREVENT trial, in particular Prof Lekander and his group at Karolinska Institutet Sweden who contributed with critical input on the biomarker assessments, and our user representative, Tove Aae, who contributed with invaluable input throughout the planning of the project.

## Availability of data and materials

An anonymized dataset of the quantitative part of the study will be made available upon reasonable request, and in line with Norwegian privacy legislation, when the main results of the trial are published. Due to privacy concerns, the qualitative data will not be shared.

## Authors’ contributions

SER is the principal investigator of the study, HBJ is the co-PI of the study. AM is the trial coordinator, while MTSH is responsible for the process evaluation. RSF is the trial statistician. SER and HBJ drafted the manuscript, with critical intellectual input from AM and MTSH. SER is responsible for the PRO content of the trial protocol. All authors read and approved the final manuscript.

## Consent for publication

Not applicable.

## Ethics approval and consent to participate

The study will comply with good clinical practice, including the most recent version of the declaration of Helsinki, as well as all relevant rules and regulations of Norway. The study has been approved by the Regional ethical committee (reference number 67725), the Data Protection Officer at Oslo University Hospital (reference number 20/00831), and the Norwegian Social Science Data Services (project number: 219790). We follow the institutional guidelines for collecting and storing sensitive data, as well as registration of the project in the University’s research data base (FORSKPRO). Prior to inclusion, all participants receive oral as well as written information about the trial, the randomization process, personal confidentiality, and the right to withdraw from the project at any time. They also sign declarations of informed consent and voluntary participation. Important protocol modifications will be reported to the ethical committee as well as the data protection officers.

## References

1. McPherson K, Steel CM, Dixon JM. ABC of breast diseases. Breast cancer-epidemiology, risk factors, and genetics. BMJ. 2000;321(7261):624-8.

2. Mouridsen HT, Bjerre KD, Christiansen P, Jensen MB, Moller S. Improvement of prognosis in breast cancer in Denmark 1977-2006, based on the nationwide reporting to the DBCG Registry. Acta Oncol. 2008;47(4):525–36.

3. Gartner R, Jensen MB, Nielsen J, Ewertz M, Kroman N, Kehlet H. Prevalence of and factors associated with persistent pain following breast cancer surgery. JAMA. 2009;302(18):1985–92.

4. Peuckmann V, Ekholm O, Rasmussen NK, Groenvold M, Christiansen P, Moller S, et al. Chronic pain and other sequelae in long-term breast cancer survivors: nationwide survey in Denmark. Eur J Pain. 2009;13(5):478–85.

5. Macdonald L, Bruce J, Scott NW, Smith WC, Chambers WA. Long-term follow-up of breast cancer survivors with post-mastectomy pain syndrome. Br J Cancer. 2005;92(2):225–30.

6. Andersen KG, Kehlet H. Persistent pain after breast cancer treatment: a critical review of risk factors and strategies for prevention. J Pain. 2011;12(7):725–46.

7. Schou Bredal I, Smeby NA, Ottesen S, Warncke T, Schlichting E. Chronic pain in breast cancer survivors: comparison of psychosocial, surgical, and medical characteristics between survivors with and without pain. J Pain Symptom Manage. 2014;48(5):852–62.

8. Mejdahl MK, Andersen KG, Gartner R, Kroman N, Kehlet H. Persistent pain and sensory disturbances after treatment for breast cancer: six year nationwide follow-up study. BMJ. 2013;346:f1865.

9. Boehmke MM, Dickerson SS. Symptom, symptom experiences, and symptom distress encountered by women with breast cancer undergoing current treatment modalities. Cancer Nurs. 2005;28(5):382–9.

10. Reinertsen KV, Loge JH, Brekke M, Kiserud CE. Chronic fatigue in adult cancer survivors. Tidsskrift for den Norske laegeforening : tidsskrift for praktisk medicin, ny raekke. 2017;137(21).

11. Chalder T, Berelowitz G, Pawlikowska T, Watts L, Wessely S, Wright D, et al. Development of a fatigue scale. Journal of Psychosomatic Research. 1993;37(2):147–53.

12. Huang HP, Chen ML, Liang J, Miaskowski C. Changes in and predictors of severity of fatigue in women with breast cancer: A longitudinal study. Int J Nurs Stud. 2014;51(4):582–92.

13. Bower JE, Ganz PA, Desmond KA, Rowland JH, Meyerowitz BE, Belin TR. Fatigue in breast cancer survivors: occurrence, correlates, and impact on quality of life. J Clin Oncol. 2000;18(4):743–53.

14. Crombie IK, Davies HT, Macrae WA. Cut and thrust: antecedent surgery and trauma among patients attending a chronic pain clinic. Pain. 1998;76(1-2):167–71.

15. Treede RD, Rief W, Barke A, Aziz Q, Bennett MI, Benoliel R, et al. A classification of chronic pain for ICD-11. Pain. 2015;156(6):1003–7.

16. Macrae WA. Chronic post-surgical pain: 10 years on. Br J Anaesth. 2008;101(1):77–86.

17. Werner MU, Kongsgaard UE. I. Defining persistent post-surgical pain: is an update required? Br J Anaesth. 2014;113(1):1–4.

18. Kehlet H, Jensen TS, Woolf CJ. Persistent postsurgical pain: risk factors and prevention. Lancet. 2006;367(9522):1618-25.

19. Munk A, Reme SE, Jacobsen HB. What Does CATS Have to Do With Cancer? The Cognitive Activation Theory of Stress (CATS) Forms the SURGE Model of Chronic Post-surgical Pain in Women With Breast Cancer. Frontiers in Psychology. 2021;12:872.

20. Hinrichs-Rocker A, Schulz K, Jarvinen I, Lefering R, Simanski C, Neugebauer EA. Psychosocial predictors and correlates for chronic post-surgical pain (CPSP) - a systematic review. Eur J Pain. 2009;13(7):719–30.

21. VanDenKerkhof EG, Hopman WM, Goldstein DH, Wilson RA, Towheed TE, Lam M, et al. Impact of perioperative pain intensity, pain qualities, and opioid use on chronic pain after surgery: a prospective cohort study. Reg Anesth Pain Med. 2012;37(1):19–27.

22. Montgomery GH, Hallquist MN, Schnur JB, David D, Silverstein JH, Bovbjerg DH. Mediators of a brief hypnosis intervention to control side effects in breast surgery patients: response expectancies and emotional distress. J Consult Clin Psychol. 2010;78(1):80–8.

23. Wang L, Guyatt GH, Kennedy SA, Romerosa B, Kwon HY, Kaushal A, et al. Predictors of persistent pain after breast cancer surgery: a systematic review and meta-analysis of observational studies. CMAJ. 2016;188(14):E352–E61.

24. De Vries J, Van der Steeg AF, Roukema JA. Determinants of fatigue 6 and 12 months after surgery in women with early-stage breast cancer: a comparison with women with benign breast problems. J Psychosom Res. 2009;66(6):495–502.

25. De Kock M. Expanding our horizons: transition of acute postoperative pain to persistent pain and establishment of chronic postsurgical pain services. Anesthesiology. 2009;111(3):461–3.

26. Meretoja TJ, Andersen KG, Bruce J, Haasio L, Sipila R, Scott NW, et al. Clinical Prediction Model and Tool for Assessing Risk of Persistent Pain After Breast Cancer Surgery. J Clin Oncol. 2017;35(15):1660–7.

27. Pinto PR, McIntyre T, Araujo-Soares V, Costa P, Almeida A. Differential predictors of acute post-surgical pain intensity after abdominal hysterectomy and major joint arthroplasty. Ann Behav Med. 2015;49(3):384–97.

28. Theunissen M, Peters ML, Bruce J, Gramke HF, Marcus MA. Preoperative anxiety and catastrophizing: a systematic review and meta-analysis of the association with chronic postsurgical pain. Clin J Pain. 2012;28(9):819–41.

29. Ursin H, Eriksen HR. The cognitive activation theory of stress. Psychoneuroendocrinology. 2004;29(5):567–92.

30. Barrett LF, Simmons WK. Interoceptive predictions in the brain. Nat Rev Neurosci. 2015;16(7):419–29.

31. Lynn SJ, Martin DJ, Frauman DC. Does hypnosis pose special risks for negative effects? A master class commentary. The International journal of clinical and experimental hypnosis. 1996;44(1):7–19.

32. Montgomery GH, David D, Winkel G, Silverstein JH, Bovbjerg DH. The effectiveness of adjunctive hypnosis with surgical patients: a meta-analysis. Anesth Analg. 2002;94(6):1639–45, table of contents.

33. Montgomery GH, DuHamel KN, Redd WH. A meta-analysis of hypnotically induced analgesia: how effective is hypnosis? The International journal of clinical and experimental hypnosis. 2000;48(2):138–53.

34. Redd WH, Montgomery GH, DuHamel KN. Behavioral intervention for cancer treatment side effects. J Natl Cancer Inst. 2001;93(11):810–23.

35. Patterson DR, Hoffman HG, Palacios AG, Jensen MJ. Analgesic effects of posthypnotic suggestions and virtual reality distraction on thermal pain. J Abnorm Psychol. 2006;115(4):834–41.

36. Enqvist B, Bjorklund C, Engman M, Jakobsson J. Preoperative hypnosis reduces postoperative vomiting after surgery of the breasts. A prospective, randomized and blinded study. Acta Anaesthesiol Scand. 1997;41(8):1028–32.

37. Faymonville ME, Fissette J, Mambourg PH, Roediger L, Joris J, Lamy M. Hypnosis as adjunct therapy in conscious sedation for plastic surgery. Reg Anesth. 1995;20(2):145–51.

38. Lang EV, Berbaum KS, Faintuch S, Hatsiopoulou O, Halsey N, Li X, et al. Adjunctive self-hypnotic relaxation for outpatient medical procedures: a prospective randomized trial with women undergoing large core breast biopsy. Pain. 2006;126(1-3):155–64.

39. Spiegel D, Bloom JR. Group therapy and hypnosis reduce metastatic breast carcinoma pain. Psychosom Med. 1983;45(4):333–9.

40. Montgomery GH, Bovbjerg DH, Schnur JB, David D, Goldfarb A, Weltz CR, et al. A randomized clinical trial of a brief hypnosis intervention to control side effects in breast surgery patients. J Natl Cancer Inst. 2007;99(17):1304–12.

41. Amraoui J, Pouliquen C, Fraisse J, Dubourdieu J, Rey Dit Guzer S, Leclerc G, et al. Effects of a Hypnosis Session Before General Anesthesia on Postoperative Outcomes in Patients Who Underwent Minor Breast Cancer Surgery: The HYPNOSEIN Randomized Clinical Trial. JAMA Netw Open. 2018;1(4):e181164.

42. Ost LG. The efficacy of Acceptance and Commitment Therapy: an updated systematic review and meta-analysis. Behav Res Ther. 2014;61:105–21.

43. McCracken LM, Morley S. The psychological flexibility model: a basis for integration and progress in psychological approaches to chronic pain management. J Pain. 2014;15(3):221–34.

44. Jacobsen HB, Kallestad H, Landro NI, Borchgrevink PC, Stiles TC. Processes in acceptance and commitment therapy and the rehabilitation of chronic fatigue. Scand J Psychol. 2017;58(3):211–20.

45. Ramsey SE, Rounsaville D, Hoskinson R, Park TW, Ames EG, Neirinckx VD, et al. The Need for Psychosocial Interventions to Facilitate the Transition to Extended-Release Naltrexone (XR-NTX) Treatment for Opioid Dependence: A Concise Review of the Literature. Subst Abuse. 2016;10:65–8.

46. Hayes SC, Luoma JB, Bond FW, Masuda A, Lillis J. Acceptance and commitment therapy: model, processes and outcomes. Behav Res Ther. 2006;44(1):1–25.

47. Weinrib AZ, Azam MA, Birnie KA, Burns LC, Clarke H, Katz J. The psychology of chronic post-surgical pain: new frontiers in risk factor identification, prevention and management. Br J Pain. 2017;11(4):169–77.

48. Katz J, Weinrib A, Fashler SR, Katznelzon R, Shah BR, Ladak SS, et al. The Toronto General Hospital Transitional Pain Service: development and implementation of a multidisciplinary program to prevent chronic postsurgical pain. Journal of pain research. 2015;8:695–702.

49. Abid Azam M, Weinrib AZ, Montbriand J, Burns LC, McMillan K, Clarke H, et al. Acceptance and Commitment Therapy to manage pain and opioid use after major surgery: Preliminary outcomes from the Toronto General Hospital Transitional Pain Service. Canadian Journal of Pain. 2017;1(1):37–49.

50. Johansen V, Fimland MS. Sluttrapport for prosjektet ved Hysnes Helsefort. Trondheim: NTNU; 2016.

51. Hara KW, Bjorngaard JH, Brage S, Borchgrevink PC, Halsteinli V, Stiles TC, et al. Randomized Controlled Trial of Adding Telephone Follow-Up to an Occupational Rehabilitation Program to Increase Work Participation. J Occup Rehabil. 2017.

52. Ganz PA, Bower JE. Cancer related fatigue: a focus on breast cancer and Hodgkin’s disease survivors. Acta Oncol. 2007;46(4):474–9.

53. Hawker GA, Mian S, Kendzerska T, French M. Measures of adult pain: Visual Analog Scale for Pain (VAS Pain), Numeric Rating Scale for Pain (NRS Pain), McGill Pain Questionnaire (MPQ), Short-Form McGill Pain Questionnaire (SF-MPQ), Chronic Pain Grade Scale (CPGS), Short Form-36 Bodily Pain Scale (SF-36 BPS), and Measure of Intermittent and Constant Osteoarthritis Pain (ICOAP). Arthritis Care Res (Hoboken). 2011;63 Suppl 11:S240-52.

54. Breivik H, Borchgrevink PC, Allen SM, Rosseland LA, Romundstad L, Hals EK, et al. Assessment of pain. British Journal of Anaesthesia. 2008;101(1):17–24.

55. Jensen MP. The validity and reliability of pain measures in adults with cancer. J Pain. 2003;4(1):2–21.

56. Fecho K, Miller NR, Merritt SA, Klauber-Demore N, Hultman CS, Blau WS. Acute and persistent postoperative pain after breast surgery. Pain Med. 2009;10(4):708–15.

57. Meretoja TJ, Leidenius MHK, Tasmuth T, Sipila R, Kalso E. Pain at 12 months after surgery for breast cancer. JAMA. 2014;311(1):90–2.

58. Yellen SB, Cella DF, Webster K, Blendowski C, Kaplan E. Measuring fatigue and other anemia-related symptoms with the Functional Assessment of Cancer Therapy (FACT) measurement system. J Pain Symptom Manage. 1997;13(2):63–74.

59. Butt Z, Lai JS, Rao D, Heinemann AW, Bill A, Cella D. Measurement of fatigue in cancer, stroke, and HIV using the Functional Assessment of Chronic Illness Therapy - Fatigue (FACIT-F) scale. J Psychosom Res. 2013;74(1):64–8.

60. Montgomery GH, D. D, Kangas M, Green S, Sucala M, Bovbjerg DH, et al. Randomized Controlled Trial of a Cognitive-Behavioral Therapy Plus Hypnosis Intervention to Control Fatigue in Patients Undergoing Radiotherapy for Breast Cancer. Journal of Clinical Oncology. 2014;32(6):557–63.

61. Sprangers MA, Groenvold M, Arraras JI, Franklin J, te Velde A, Muller M, et al. The European Organization for Research and Treatment of Cancer breast cancer-specific quality-of-life questionnaire module: first results from a three-country field study. J Clin Oncol. 1996;14(10):2756–68.

62. Zigmond AS, Snaith RP. The hospital anxiety and depression scale. Acta Psychiatr Scand. 1983;67(6):361–70.

63. Bjelland I, Dahl AA, Haug TT, Neckelmann D. The validity of the Hospital Anxiety and Depression Scale: An updated literature review. Journal of Psychosomatic Research. 2002;52(2):69–77.

64. Wu Y, Levis B, Sun Y, He C, Krishnan A, Neupane D, et al. Accuracy of the Hospital Anxiety and Depression Scale Depression subscale (HADS-D) to screen for major depression: systematic review and individual participant data meta-analysis. Bmj. 2021;373:n972.

65. Guy W. ECDEU Assessment Manual for Psychopharmacology, Revised. Rockville: National Institute of Mental Health; 1976.

66. Dworkin RH, Turk DC, Wyrwich KW, Beaton D, Cleeland CS, Farrar JT, et al. Interpreting the clinical importance of treatment outcomes in chronic pain clinical trials: IMMPACT recommendations. The Journal of Pain. 2008;9(2):105–21.

67. Perrot S, Lantéri-Minet M. Patients’ Global Impression of Change in the management of peripheral neuropathic pain: Clinical relevance and correlations in daily practice. Eur J Pain. 2019;23(6):1117–28.

68. Bond FW, Hayes SC, Baer RA, Carpenter KM, Guenole N, Orcutt HK, et al. Preliminary psychometric properties of the Acceptance and Action Questionnaire-II: a revised measure of psychological inflexibility and experiential avoidance. Behav Ther. 2011;42(4):676–88.

69. Shari NI, Zainal NZ, Guan NC, Ahmad Sabki Z, Yahaya NA. Psychometric properties of the acceptance and action questionnaire (AAQ II) Malay version in cancer patients. PloS one. 2019;14(2):e0212788-e.

70. Østergaard T, Lundgren T, Zettle RD, Landrø NI, Haaland VØ. Norwegian Acceptance and Action Questionnaire (NAAQ): A psychometric evaluation. Journal of Contextual Behavioral Science. 2020;15:103–9.

71. Due P, Holstein B, Lund R, Modvig J, Avlund K. Social relations: network, support and relational strain. Soc Sci Med. 1999;48(5):661–73.

72. Skovbjerg S, Rasmussen A, Zachariae R, Schmidt L, Lund R, Elberling J. The association between idiopathic environmental intolerance and psychological distress, and the influence of social support and recent major life events. Environmental Health and Preventive Medicine. 2012;17(1):2–9.

73. Sullivan MJ, Adams H, Horan S, Maher D, Boland D, Gross R. The role of perceived injustice in the experience of chronic pain and disability: scale development and validation. Journal of Occupational Rehabilitation. 2008;18(3):249–61.

74. Ljosaa TM, Berg HS, Jacobsen HB, Granan L-P, Reme S. Translation and validation of the Norwegian version of the Injustice Experience Questionnaire. Scandinavian Journal of Pain. 2021.

75. American Psychiatric Association. Diagnostic and Statistical Manual of Mental Disorders: DSM-IV. 4th ed. Washington, DC 1994.

76. Pallesen S, Bjorvatn B, Nordhus IH, Sivertsen B, Hjørnevik M, Morin CM. A new scale for measuring insomnia: the Bergen Insomnia Scale. Percept Mot Skills. 2008;107(3):691–706.

77. Duffy D, Rouilly V, Libri V, Hasan M, Beitz B, David M, et al. Functional analysis via standardized whole-blood stimulation systems defines the boundaries of a healthy immune response to complex stimuli. Immunity. 2014;40(3):436–50.

78. Staufenbiel SM, Penninx BW, Spijker AT, Elzinga BM, van Rossum EF. Hair cortisol, stress exposure, and mental health in humans: a systematic review. Psychoneuroendocrinology. 2013;38(8):1220–35.

79. Dettenborn L, Tietze A, Kirschbaum C, Stalder T. The assessment of cortisol in human hair: associations with sociodemographic variables and potential confounders. Stress. 2012;15(6):578–88.

80. Wolfe F, Clauw DJ, Fitzcharles M-A, Goldenberg DL, Häuser W, Katz RL, et al. 2016 Revisions to the 2010/2011 fibromyalgia diagnostic criteria. Seminars in Arthritis and Rheumatism. 2016;46(3):319-29.

81. Fors EA, Wensaas K-A, Eide H, Jaatun EA, Clauw DJ, Wolfe F, et al. Fibromyalgia 2016 criteria and assessments: comprehensive validation in a Norwegian population. Scandinavian Journal of Pain. 2020;20(4):663-72.

82. Sullivan MJ, Thorn B, Haythornthwaite JA, Keefe F, Martin M, Bradley LA, et al. Theoretical perspectives on the relation between catastrophizing and pain. Clin J Pain. 2001;17(1):52–64.

83. Bot AG, Becker SJ, van Dijk CN, Ring D, Vranceanu AM. Abbreviated psychologic questionnaires are valid in patients with hand conditions. Clin Orthop Relat Res. 2013;471(12):4037–44.

84. Walton DM, Mehta S, Seo W, MacDermid JC. Creation and validation of the 4-item BriefPCS-chronic through methodological triangulation. Health and Quality of Life Outcomes. 2020;18(1):124.

85. Pan PH, Tonidandel AM, Aschenbrenner CA, Houle TT, Harris LC, Eisenach JC. Predicting acute pain after cesarean delivery using three simple questions. Anesthesiology. 2013;118(5):1170–9.

86. Lovvik C, Shaw W, Overland S, Reme SE. Expectations and illness perceptions as predictors of benefit recipiency among workers with common mental disorders: secondary analysis from a randomised controlled trial. BMJ Open. 2014;4(3):e004321.

87. Lovvik C, Overland S, Hysing M, Broadbent E, Reme SE. Association between illness perceptions and return-to-work expectations in workers with common mental health symptoms. J Occup Rehabil. 2014;24(1):160–70.

88. Reme SE, Hagen EM, Eriksen HR. Expectations, perceptions, and physiotherapy predict prolonged sick leave in subacute low back pain. BMC Musculoskelet Disord. 2009;10(1):139.

89. Reme SE, Grasdal AL, Lovvik C, Lie SA, Overland S. Work-focused cognitive-behavioural therapy and individual job support to increase work participation in common mental disorders: a randomised controlled multicentre trial. Occup Environ Med. 2015;72(10):745–52.

90. Theunissen M, Peters ML, Schouten EG, Fiddelers AA, Willemsen MG, Pinto PR, et al. Validation of the surgical fear questionnaire in adult patients waiting for elective surgery. PLoS One. 2014;9(6):e100225.

91. Hinz A, Sander C, Glaesmer H, Brahler E, Zenger M, Hilbert A, et al. Optimism and pessimism in the general population: Psychometric properties of the Life Orientation Test (LOT-R). Int J Clin Health Psychol. 2017;17(2):161–70.

92. Schou-Bredal I, Heir T, Skogstad L, Bonsaksen T, Lerdal A, Grimholt T, et al. Population-based norms of the Life Orientation Test–Revised (LOT-R). International Journal of Clinical and Health Psychology. 2017;17(3):216–24.

93. Data management plan [Internet]. 2022. Available from: https://www.uio.no/for-ansatte/arbeidsstotte/forskningsstotte/forskpro/prosjekter/sv/psi/pre--and-post-operative-psychological-interventions-to-prevent-pain-and-fatigue-after-breast-cancer-surgery-a-randomized-controlled-trial/index.html.

94. Lind SB, Jacobsen HB, Solbakken OA, Reme SE. Clinical Hypnosis in Medical Care: A Mixed-Method Feasibility Study. Integrative Cancer Therapies. 2021;20:15347354211058678.

95. Veehof MM, Trompetter HR, Bohlmeijer ET, Schreurs KM. Acceptance- and mindfulness-based interventions for the treatment of chronic pain: a meta-analytic review. Cogn Behav Ther. 2016;45(1):5–31.

96. Strosahl KD, Robinson PJ, Gustavsson T. Brief interventions for radical change: Principles and practice of focused acceptance and commitment therapy: New Harbinger Publications; 2012.

97. Trompetter HR, Bohlmeijer ET, Veehof MM, Schreurs KM. Internet-based guided self-help intervention for chronic pain based on Acceptance and Commitment Therapy: a randomized controlled trial. J Behav Med. 2015;38(1):66–80.

98. Bewick BM, Ondersma SJ, Hoybye MT, Blakstad O, Blankers M, Brendryen H, et al. Key Intervention Characteristics in e-Health: Steps Towards Standardized Communication. International journal of behavioral medicine. 2017;24(5):659–64.

99. Danaher BG, Brendryen H, Seeley JR, Tyler MS, Woolley T. From black box to toolbox: Outlining device functionality, engagement activities, and the pervasive information architecture of mHealth interventions. Internet Interv. 2015;2(1):91–101.

100. Montgomery GH, Kangas M, David D, Hallquist MN, Green S, Bovbjerg DH, et al. Fatigue during breast cancer radiotherapy: an initial randomized study of cognitive-behavioral therapy plus hypnosis. Health Psychol. 2009;28(3):317–22.

101. Johannsen M, O’Connor M, O’Toole MS, Jensen AB, Hojris I, Zachariae R. Efficacy of Mindfulness-Based Cognitive Therapy on Late Post-Treatment Pain in Women Treated for Primary Breast Cancer: A Randomized Controlled Trial. J Clin Oncol. 2016;34(28):3390–9.

102. Jackson DL. Reporting results of latent growth modeling and multilevel modeling analyses: some recommendations for rehabilitation psychology. Rehabilitation psychology. 2010;55(3):272–85.

103. Rabe-Hesketh S, A. S. Multilevel and Longitudinal Modeling Using Stata. Texas: Stata Press; 2008.

104. Brinkmann S, Kvale S. InterViews: Learning the Craft of Qualitative Interviewing. Third edition ed: SAGE publications Inc; 2015.

105. Polkinghorne DE. Language and meaning: Data collection in qualitative research. Journal of Counseling Psychology. 2005;52(2):137–45.

106. Frank A. Letting stories breathe: A socio-narratology.: The University of Chicago Press; 2012.

107. Riessmann C. Narrative methods for the human sciences. Thousand Oaks, CA. : SAGE Publications, Inc. ; 2008.

108. Charmaz K. Constructing Grounded Theory: A practical guide through qualitative analysis.: SAGE Publications, Inc.; 2014.

109. Levitt HM, Bamberg M, Creswell JW, Frost DM, Josselson R, Suarez-Orozco C. Journal article reporting standards for qualitative primary, qualitative meta-analytic, and mixed methods research in psychology: The APA Publications and Communications Board task force report. Am Psychol. 2018;73(1):26–46.

110. Birch S, Stilling M, Mechlenburg I, Hansen TB. Effectiveness of a physiotherapist delivered cognitive-behavioral patient education for patients who undergoes operation for total knee arthroplasty: a protocol of a randomized controlled trial. BMC musculoskeletal disorders. 2017;18(1):116.

